# Low-Frequency Activity in Dorsal Subthalamic Nucleus Predicts Impulsivity Improvement Following Deep Brain Stimulation in Parkinsonian Patients

**DOI:** 10.1101/2024.05.10.24307166

**Authors:** Ahmet Kaymak, Laura Cubeddu, Matteo Vissani, Fabio Taddeini, Luca Caremani, Alessandra Govoni, Federico Micheli, Simone Valente, Francesca Piattellini, Davide Greco, Guido Pecchioli, Silvia Ramat, Alberto Mazzoni

## Abstract

**Objective:** The progression of impulsive-compulsive behaviors (ICB) in Parkinson’s Disease (PD) following subthalamic deep brain stimulation (DBS) surgery displays a large inter-patient variability. However, the link between the subthalamic neural activity at the single-neuron level and the postoperative evolution of ICB remains unclear. In this study, we investigated neural features associated with postoperative ICB recovery and their spatial distribution within the subthalamic nucleus (STN).

**Approach:** We examined neural activity extracted from intraoperative microelectrode recordings within the STN of 22 PD patients undergoing STN-DBS. Ten patients were diagnosed with ICB, with half of them showing recovery (ICB-R) from impulsive symptoms following implantation, while the other half remained stable (ICB-S). Both groups presented similar motor symptoms and received similar drug treatments pre- and post-operatively. Following, we compared beta [12-30 Hz] and theta [4-8 Hz] oscillations, firing rate, regularity, and spiking patterns in non-ICB, recovered, and stable patients across STN regions. We adopted linear discriminant algorithms to classify the postoperative state at both single neuron and patient levels.

**Main results:** We observed significantly weaker beta and theta oscillations and increased spiking regularity at the single neuron level (*p*<0.05, Mann-Whitney U test) in patients who displayed postoperative ICB recovery. Of note, this difference was significant only on the dorsal portion of the STN, close to the stimulation target region. The discrimination algorithms based on these features correctly classified the postoperative state of 9/10 ICB patients.

**Significance:** We showed that low-frequency subthalamic neural activity next to the stimulation target could be an effective biomarker for the evolution of ICBs following STN-DBS surgery, independently from other clinical aspects. Our results also support broader implications of beta activity in PD pathology beyond the motor domain.

## 1. Introduction

Parkinson’s disease (PD) is a neurodegenerative condition characterized by motor dysfunction and both cognitive and psychological disturbances.^1^ PD can lead to behavioral problems related to impulse control disorders (ICBs) in 14 to 40%^2^ of the subjects.^1^

The development of impulsive behaviors in PD patients is associated with multiple factors. Primarily, dopamine replacement therapy (DRT) has been linked to a 2 to 7-fold increased risk^3^ of developing ICBs for patients and these symptoms usually improve with the reduction of DRT treatment.^4^ The heightened risk is believed to stem from the overactivation of the mesolimbic dopaminergic system, which is involved in modulating behavioral responses linked to reward, motivation, and reinforcement.^5^ Impulsivity may also manifest in PD patients who are subjected to higher levodopa intake.^6^ Additionally, non-dopaminergic factors such as younger PD onset, male gender, unmarried status, and history of alcohol can also display a significant correlation with ICBs symptoms.^6,7^

The impact of subthalamic nucleus (STN) deep brain stimulation (DBS) on impulsivity symptoms remains uncertain.^8^ Direct and indirect mechanisms have been proposed to explain the efficacy of STN-DBS surgery for alleviating ICBs symptoms in PD patients.^9–13^ Indirectly, STN-DBS alleviates motor complaints following the intervention and reduces the dependency on dopaminergic medication. This reduction in medication use can lead to improvements in impulsivity.^9,11,12^ Considering the role of the median limbic part of STN in cognition and emotion^14^ and the involvement of STN in the brain’s reward neural pathways^15^, subthalamic stimulation can also directly act on circuits underlying ICB. Coherently, the stimulation position within the STN was found to impact the postoperative neuropsychiatric symptoms in PD patients.^13,16^ Overall, factors including preoperative clinical conditions, surgical methods, and DBS targeting contribute to the variation in postoperative impulsivity improvement.^7,17^

Multiple studies have investigated separately the preoperative^10,18^ and DBS targeting^13,16^ associated with the postoperative improvement of ICBs. However, not all patients experience improvement in impulsivity following STN-DBS, and the connections between subthalamic electrophysiology at the single neuron level and preoperative clinical factors in improved and non-improved cases remain to be elucidated. Our research examined disparities between two patient groups regarding their subthalamic neural activity and observed the spatial behaviors of neural patterns associated with the postoperative evolution of impulsive behaviors. Lastly, we demonstrated the utility of simple linear algorithms that incorporate subthalamic neural activity in estimating the likelihood of impulsivity improvement in PD patients.

## 2. Methods

### 2.1 Patient Selection

The study is a retrospective cohort study conducted under the Declaration of Helsinki and after ethical committee approval (Careggi Hospital, Florence, Italy). For full details regarding the diagnosis criteria for PD and ICBs, patient selection, and surgical and electrophysiological recording techniques we refer to two previous works.^19,20^ Briefly, the PD diagnosis was made based on the UK Parkinson’s Disease Society Brain Bank diagnostic criteria at the preoperative baseline.^21^ For this study, we focused on PD patients undergoing STN-DBS and presenting at least one current ICB accompanying motor symptoms (ICB)(n=12). Among ICB patients, we identified two groups: those who exhibited recovery in impulsive behavior (n=5) (ICB-R) following DBS, and those who remained stable regarding impulsive behaviors (n=5) (ICB-S).

To ensure a comparable preoperative baseline between the two groups and prevent confounding baseline factors that could affect neural activity (particularly motor severity which is strongly associated with beta band activity in STN^22^), we excluded 2 ICB-S patients from further analysis, a departure from two previous studies. Furthermore, we included a control group consisting of 12 age- and sex-matched non-ICB PD patients. The degree of impulsivity was measured with the Barratt Impulsiveness Scale(BIS).^23^

### 2.2 Study Design

Preoperatively, patients were subjected to dopamine replacement therapies to improve their motor symptoms. The dosages of dopamine agonist (DA) agents and dopamine replacement drugs were measured in terms of their levodopa equivalent daily doses (LEDD). The effects of pharmacological treatments were evaluated by Hoehn and Yahr^24^, and the Movement Disorders Society Unified Parkinson’s Disease Rating Scale part III (UPDRS III)^25^ scales under medication on and off conditions. Subsequently, we noted clinical factors including the duration of PD, the severity of motor and cognitive symptoms, along with pharmacological treatment and demographic information (Figure 1a, Preoperative Phase).

**Figure 1.**
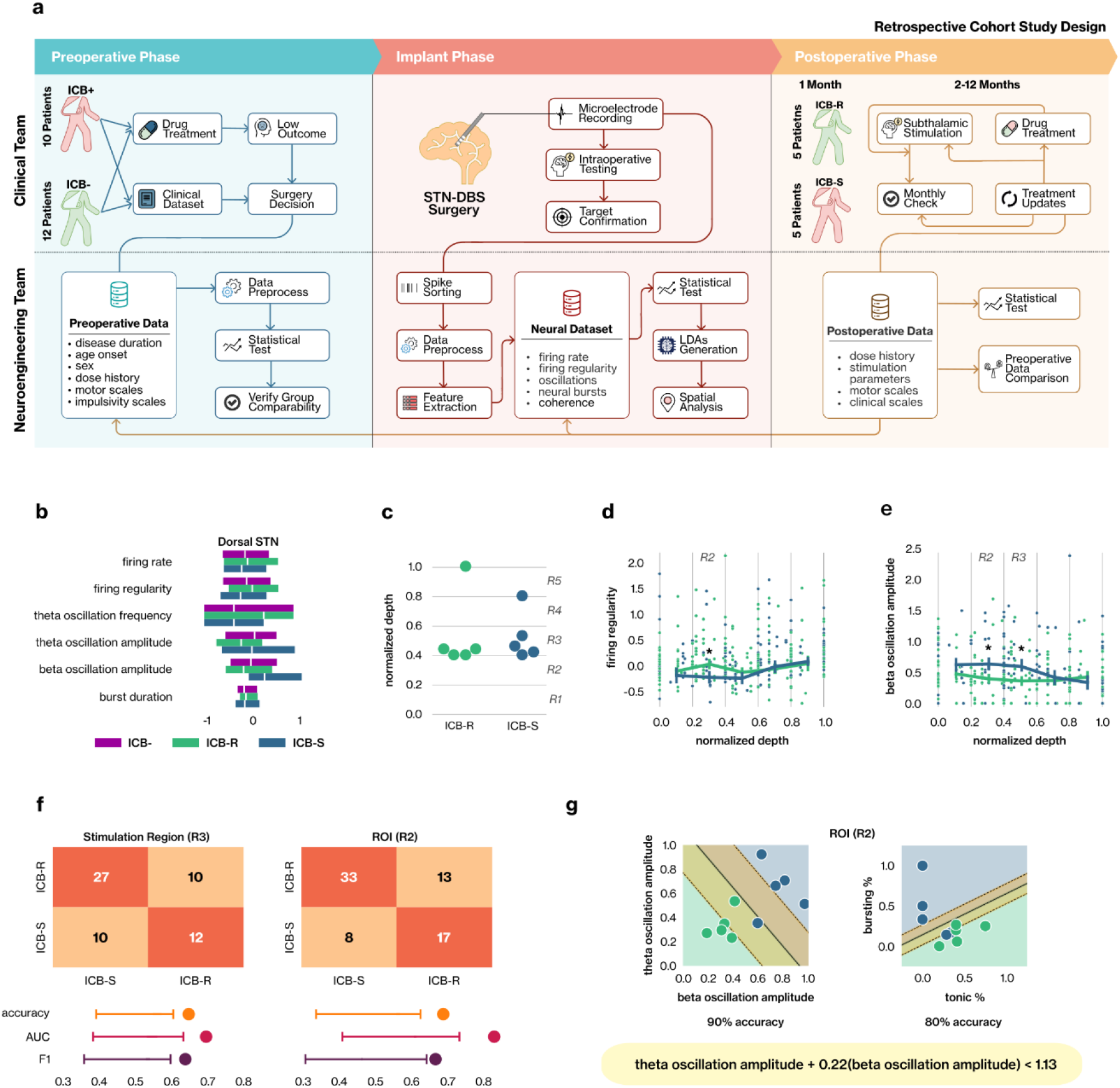
Cohort Study Design & Neural Analysis. **(a)** Retrospective cohort study design. Clinical and neural data are collected in three different phases (columns from left to right: preoperative, during the implant, and postoperative). The top row illustrates the data acquisition performed by the clinical team, and the bottom row the data analysis performed by the neuroengineering team. **(b)** The value distribution of neural features (z-scores) among ICB-R (green), ICB-S (blue), and ICB-(purple) patients in the dorsal STN is shown. The median value is depicted by the white line, and the interquartile range is represented by the edges of the bars for each feature. **(c)** The positions of the stimulation target on normalized depths along the dorsoventral axis of the STN were indicated between ICB-R and ICB-S patients. **(d, e)** The spatial characteristics of firing regularity (d) and beta oscillation amplitude (e) along the normalized depth of the dorsoventral axis of the STN are illustrated. The scatter plot shows the position of the neuron and its corresponding value of the selected neural feature in two-dimensional space. The line plot depicts the median values of the selected features in five spatial regions. Statistical comparisons revealing significant effects between ICB-S and ICB-R patients in specific regions are denoted with a star (*: *P*≤0.05). **(f)** The decoding performances of subthalamic neurons between the two groups are presented for R3 (left) and R2 (right) regions. The confusion matrix illustrates the overall classification capability of the trained SVM model with stratified 5-fold cross-validation. Average performances (across stratified 5-folds) of neuron classification are denoted with dots, and corresponding random chance intervals with upper and lower bounds are indicated with vertical lines. **(g)** Linear discrimination between ICB-S and ICB-R subjects is depicted with single points representing the combined median values of selected neural features in two-dimensional space. The linear median decision boundary is represented by a black line, and the margin of the hyperplane is shown as an orange area, determined through linear support vector machines.

Following baseline assessment, the patients underwent STN-DBS surgery where explorative microelectrode recordings (MERs) were used to detect the optimal DBS target. Then, we characterized the subthalamic neural activity regarding firing rate, firing regularity, and pathological oscillations in beta and theta bands.^20^ A comparative analysis was carried out between ICB-R and ICB-S patients to identify neural features that are strongly associated with postoperative ICBs improvement. We additionally investigated the spatial characteristics of these features within STN (Figure 1, Implant Phase).

In the postoperative phase, patients underwent regular monthly evaluations to monitor their progress. The stimulation parameters and pharmacological treatments were adjusted as needed to optimize the treatment outcomes throughout their recovery process (Figure 1, Postoperative Phase). Monthly visits both included clinical examination and a semi-structured interview about the presence of clinically significant compulsive buying, gambling, eating, punding, and sexual behaviors to assess actual ICBs under medication off, stimulation on condition. Subjects were asked to self-rate whether their ICD disorder was in full remission, partial remission, or remained fully symptomatic. In the monitoring period, we observed that some patients showed full recovery within the first month and maintained this recovery throughout the follow-up period (ICB-R group).

### 2.3 Statistical Analysis

Demographic, clinical, and neural characteristics were compared between the ICB-S, ICB-R, and non-ICB groups. Mann-Whitney U and Fisher’s exact test were conducted for numerical and categorical variables respectively. Spearman’s correlation coefficient is measured to evaluate the linear relationship between variables. To address the statistical significance of the degree of correlation, we conducted the Wald Test with t-distribution. Statistical significance is determined with *α*=0.05 and all p-values are corrected with Holm-Bonferroni correction.

### 2.4 Neural Data Analyses

#### 2.4.1 Spike Detection and Neural Feature Estimation

From MERs, we isolated 312 stable neurons for ICB (195 ICB-R and 117 ICB-S) and 330 stable neurons for non-ICB groups by using MATLAB ToolBox WaveClus^26^. For each neuron, we measured the neural features associated with firing patterns and bursting activity such as firing rate^27^ and a fraction of tonic neurons^28^. Intraburst frequency, interburst interval, and total bursting duration were measured based on the Rank surprise method^29^ and averaged over each neuron. The anatomical locations of recordings were expressed as normalized depths along the trajectory, with 0 representing the dorsal STN entrance and 1 indicating the ventral STN exit, based on electrophysiological-defined boundaries.^20^ Full details are reported in previous works.^19,20^

#### 2.4.2 Impulsivity Decoding at Neuron and Patient Levels

At the single-neuron level, we examined neural patterns between ICB-R and ICB-S patients across different spatial scales: the entire STN, as well as its dorsal (depth_MER_<0.5) and ventral (depth_MER_≥0.5) segments separately. To conduct a more focused spatial analysis, we partitioned the STN into 5 regions based on normalized MER depths (0: STN entry, 1: STN exit in the dorsoventral axis), with a chosen bin size of 0.2, i.e. 20% of the total STN depth (Figure 1c). For neural classification, we utilized features showing significant effects between these two groups to train and test the linear support vector machine^30^ (SVM) models using 5-fold cross-validation. Model performance was assessed using balanced accuracy, weighted area under the curve (AUC), and F1 scores, averaged across the five folds. To assess the robustness of our model’s performance, we conducted a bootstrap test with 100 iterations. In each iteration, we randomly assigned STN neurons to ICB-S or ICB-R groups while maintaining the initial ratio. A new SVM model was trained, and its performance was measured in each iteration to build the null (random chance performance) distribution. By evaluating the 95th percentiles of the null distributions, we determined the upper boundary of the random chance interval and we verified whether the performance of our trained model exceeded it.

We attempted to decode ICB-R and ICB-S patients using the same neural features, represented by median values per patient. Subsequently, we generated neural feature pairs and placed the patient groups into the two-dimensional space created by each feature pair. We then utilized linear SVM models without hyperparameter tuning to generate a linear decision boundary that optimally separates ICB-R and ICB patients. To ensure the robustness of the separation, we conducted leave-one-patient validation which involved iteratively excluding a single patient from the analysis, training the linear SVM model by using the remaining patients (n=9), and generating a decision boundary. By employing this approach, we generated 10 linear decision boundaries and then we calculated the median decision equation to serve as a predictive neural linear discrimination algorithm (LDA) for postoperative ICB improvement. Lastly, we measured the performance of these neural LDAs.

## 3. Results

### 3.1 Comparison of Preoperative and Postoperative Clinical Profiles

As stated previously, we aimed for a comparable baseline assessment between ICB-R and ICB-S patients which was confirmed by our statistical comparisons (*p*>0.05, the Mann-Whitney U Test) (Table 1). As anticipated, there were significant differences in many of the BIS subscale scores between non-ICB and ICB-R, ICB-S patients. Interestingly, motor complaints and pharmacological treatment of the ICB-S group remained comparable to those in the non-ICB patients (Table 1).

**Table 1.**
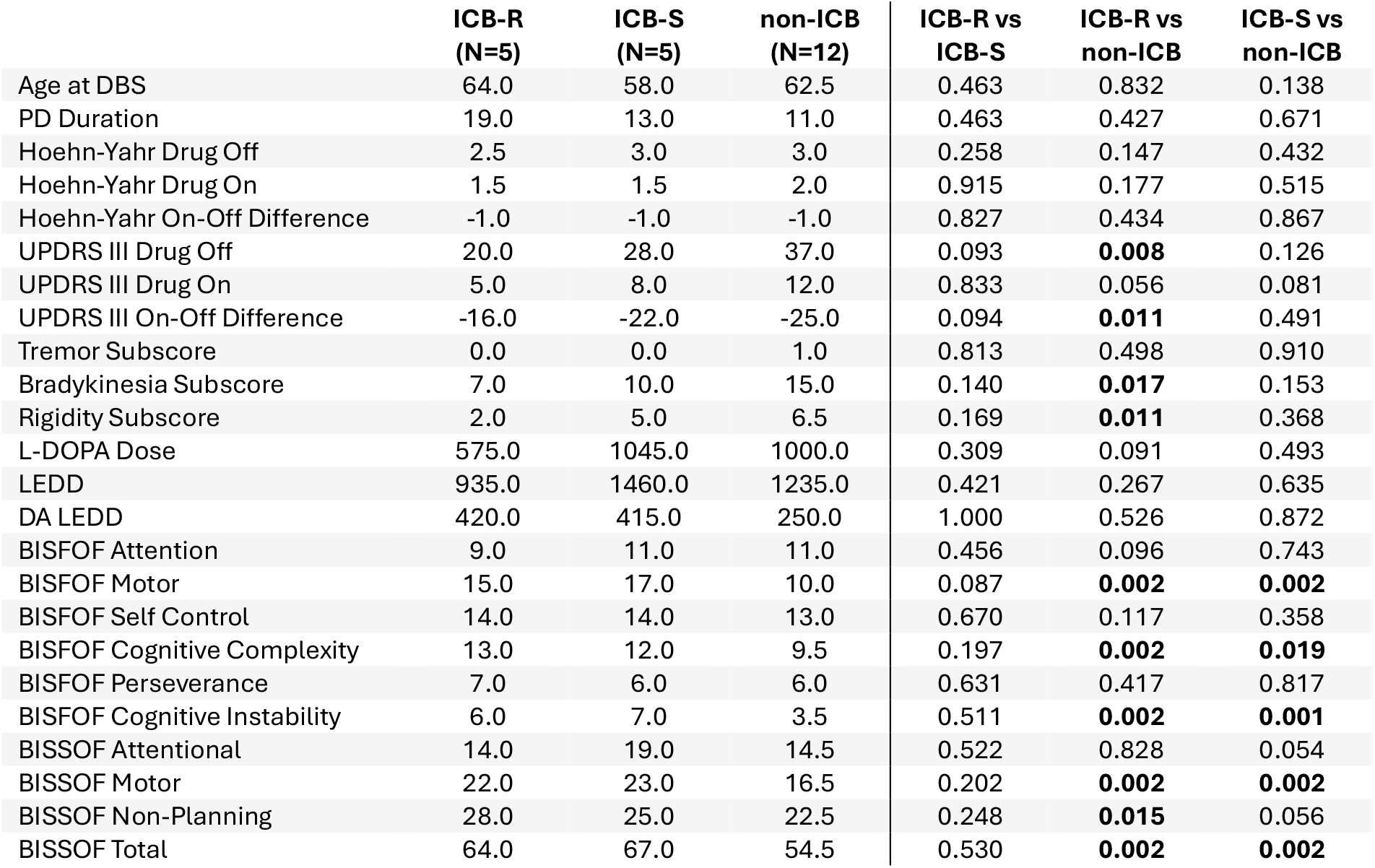
Preoperative Clinical, Demographical Characteristic, and Dopaminergic Replacement Therapy Comparison Between ICB-R, ICB-S and non-ICB Patients. The values presented in the table indicate the median values. The statistical comparison is made with the Mann-Whitney U test with the Holm-Bonferroni correction. **Abbreviations: BISFOF**, Barrat Impulsiveness Scale First Order Factor; **BISSOF**, Barrat Impulsiveness Scale Second-Order Factor; **DA**, dopamine agonist; **DBS**, deep brain stimulations; **ICB**, impulsive control behavior; **DRT**, dopaminergic replacement therapy; **LEDD**, levodopa equivalent daily doses; **PD**, Parkinson’s Disease.

|  | ICB-R<br>(N=5) | ICB-S<br>(N=5) | non-ICB<br>(N=12) | ICB-R vs<br>ICB-S | ICB-R vs<br>non-ICB | ICB-S vs<br>non-ICB |
| --- | --- | --- | --- | --- | --- | --- |
| Age at DBS | 64.0 | 58.0 | 62.5 | 0.463 | 0.832 | 0.138 |
| PD Duration | 19.0 | 13.0 | 11.0 | 0.463 | 0.427 | 0.671 |
| Hoehn-Yahr Drug Off | 2.5 | 3.0 | 3.0 | 0.258 | 0.147 | 0.432 |
| Hoehn-Yahr Drug On | 1.5 | 1.5 | 2.0 | 0.915 | 0.177 | 0.515 |
| Hoehn-Yahr On-Off Difference | -1.0 | -1.0 | -1.0 | 0.827 | 0.434 | 0.867 |
| UPDRS III Drug Off | 20.0 | 28.0 | 37.0 | 0.093 | **0.008** | 0.126 |
| UPDRS III Drug On | 5.0 | 8.0 | 12.0 | 0.833 | 0.056 | 0.081 |
| UPDRS III On-Off Difference | -16.0 | -22.0 | -25.0 | 0.094 | **0.011** | 0.491 |
| Tremor Subscore | 0.0 | 0.0 | 1.0 | 0.813 | 0.498 | 0.910 |
| Bradykinesia Subscore | 7.0 | 10.0 | 15.0 | 0.140 | **0.017** | 0.153 |
| Rigidity Subscore | 2.0 | 5.0 | 6.5 | 0.169 | **0.011** | 0.368 |
| L-DOPA Dose | 575.0 | 1045.0 | 1000.0 | 0.309 | 0.091 | 0.493 |
| LEDD | 935.0 | 1460.0 | 1235.0 | 0.421 | 0.267 | 0.635 |
| DA LEDD | 420.0 | 415.0 | 250.0 | 1.000 | 0.526 | 0.872 |
| BISFOF Attention | 9.0 | 11.0 | 11.0 | 0.456 | 0.096 | 0.743 |
| BISFOF Motor | 15.0 | 17.0 | 10.0 | 0.087 | **0.002** | **0.002** |
| BISFOF Self Control | 14.0 | 14.0 | 13.0 | 0.670 | 0.117 | 0.358 |
| BISFOF Cognitive Complexity | 13.0 | 12.0 | 9.5 | 0.197 | **0.002** | **0.019** |
| BISFOF Perseverance | 7.0 | 6.0 | 6.0 | 0.631 | 0.417 | 0.817 |
| BISFOF Cognitive Instability | 6.0 | 7.0 | 3.5 | 0.511 | **0.002** | **0.001** |
| BISSOF Attentional | 14.0 | 19.0 | 14.5 | 0.522 | 0.828 | 0.054 |
| BISSOF Motor | 22.0 | 23.0 | 16.5 | 0.202 | **0.002** | **0.002** |
| BISSOF Non-Planning | 28.0 | 25.0 | 22.5 | 0.248 | **0.015** | 0.056 |
| BISSOF Total | 64.0 | 67.0 | 54.5 | 0.530 | **0.002** | **0.002** |

In the postoperative first-year evaluation, ICB-R and ICB-S patients did not present significant differences (Supplementary Table 1, left column). Furthermore, when comparing changes from the preoperative baseline to the postoperative first-year evaluation, the two groups remained similar (Supplementary Table 1, right column).

### 3.2 The Role of Dorsal Subthalamic Activity on Impulsivity Improvement

We examined whether there were significant differences in subthalamic neural activity, during the implantation, between ICB-R and ICB-S patients and whether these patterns can be used to predict postoperative improvement in ICB. The amplitude of the beta band [12-30 Hz] and the peak frequency in the theta band [4-8 Hz] significantly differed between ICB-R and ICB-S patients in the entire STN (*p*=0.015, 0.020 respectively, Mann-Whitney U test) (Table 2). ICB-R patients exhibited significantly elevated firing regularity and firing rate compared to the non-ICB patients (*p*=0.017 and 0.036 respectively), while the ICB-S and non-ICB groups showed no significant differences. Likewise, the amplitude and duration of beta bursts were significantly higher in ICB-R patients compared to non-ICB patients (Table 2).

**Table 2.**
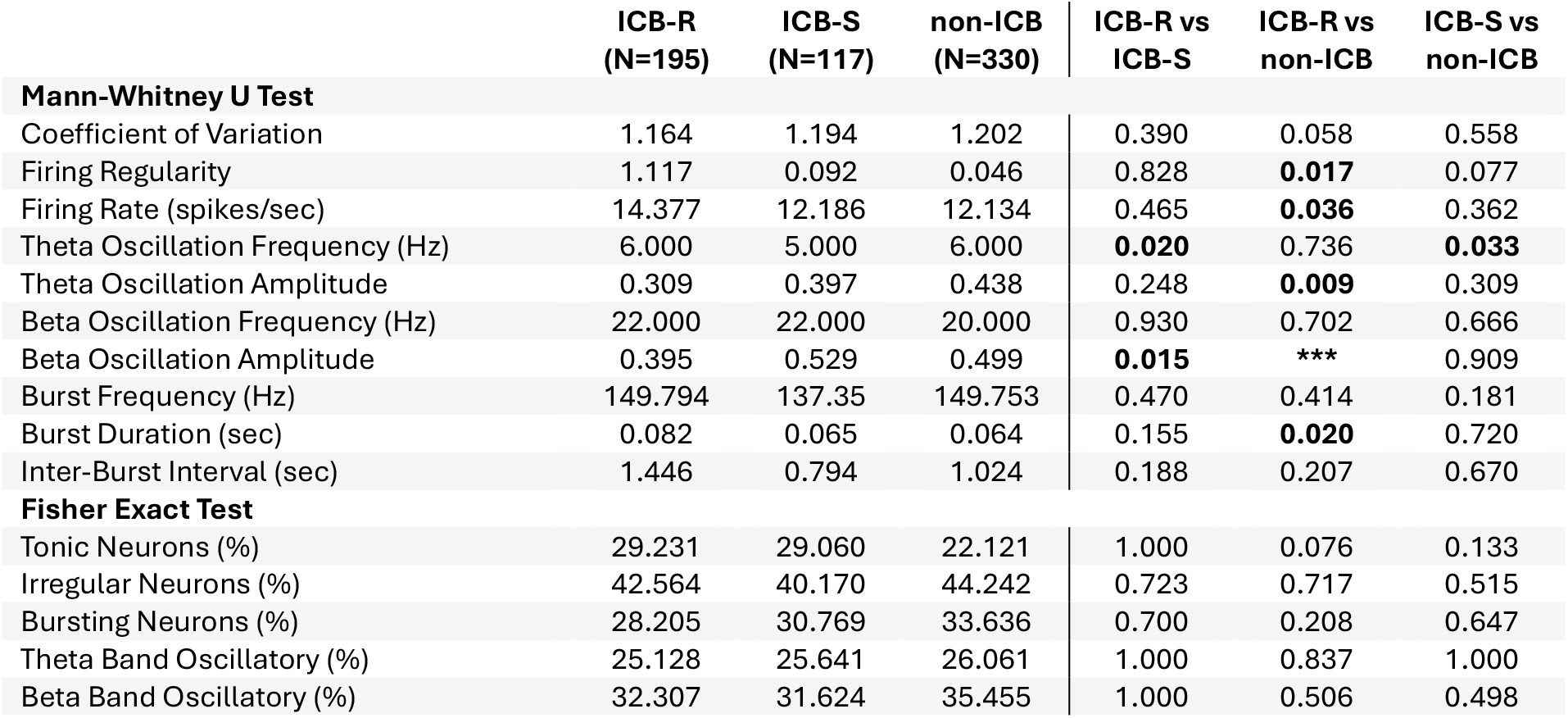
Characteristics of Subthalamic Single-Unit Activity in ICB-R vs ICB-S Patients. The statistical comparison is made with either the Mann-Whitney U test with the Holm-Bonferroni correction or the Fisher Exact test. The values presented in the table indicate the median values. **\*\*\*** *P*<0.001

|  | ICB-R<br>(N=195) | ICB-S<br>(N=117) | non-ICB<br>(N=330) | ICB-R vs<br>ICB-S | ICB-R vs<br>non-ICB | ICB-S vs<br>non-ICB |
| --- | --- | --- | --- | --- | --- | --- |
| **Mann-Whitney U Test** |  |  |  |  |  |  |
| Coefficient of Variation | 1.164 | 1.194 | 1.202 | 0.390 | 0.058 | 0.558 |
| Firing Regularity | 1.117 | 0.092 | 0.046 | 0.828 | **0.017** | 0.077 |
| Firing Rate (spikes/sec) | 14.377 | 12.186 | 12.134 | 0.465 | **0.036** | 0.362 |
| Theta Oscillation Frequency (Hz) | 6.000 | 5.000 | 6.000 | **0.020** | 0.736 | **0.033** |
| Theta Oscillation Amplitude | 0.309 | 0.397 | 0.438 | 0.248 | **0.009** | 0.309 |
| Beta Oscillation Frequency (Hz) | 22.000 | 22.000 | 20.000 | 0.930 | 0.702 | 0.666 |
| Beta Oscillation Amplitude | 0.395 | 0.529 | 0.499 | **0.015** | **\*\*\*** | 0.909 |
| Burst Frequency (Hz) | 149.794 | 137.35 | 149.753 | 0.470 | 0.414 | 0.181 |
| Burst Duration (sec) | 0.082 | 0.065 | 0.064 | 0.155 | **0.020** | 0.720 |
| Inter-Burst Interval (sec) | 1.446 | 0.794 | 1.024 | 0.188 | 0.207 | 0.670 |
| **Fisher Exact Test** |  |  |  |  |  |  |
| Tonic Neurons (%) | 29.231 | 29.060 | 22.121 | 1.000 | 0.076 | 0.133 |
| Irregular Neurons (%) | 42.564 | 40.170 | 44.242 | 0.723 | 0.717 | 0.515 |
| Bursting Neurons (%) | 28.205 | 30.769 | 33.636 | 0.700 | 0.208 | 0.647 |
| Theta Band Oscillatory (%) | 25.128 | 25.641 | 26.061 | 1.000 | 0.837 | 1.000 |
| Beta Band Oscillatory (%) | 32.307 | 31.624 | 35.455 | 1.000 | 0.506 | 0.498 |

Our team previously reported that ventral STN neurons present significantly more regular and less bursty activity for ICB patients compared to non-ICB PD patients.^19^ We investigated if a similar pattern was also evident between ICB-R (n=111) and ICB-S (n=58) neurons in ventral STN. However, the proportions of tonic and bursty neurons were not significantly different between the two groups (*p*=0.108 and 0.264 respectively, Fisher’s exact test) as well as firing regularity metric (*p*=0.185, Mann-Whitney U test) (Supplementary Table 2). These results suggest that spiking irregularity is a distinguishing factor for the existence of impulsivity in the ventral STN of PD patients, however, it is not relevant for predicting postoperative impulsivity improvement. Conversely, ICB-R patients exhibited significantly lower beta amplitude and higher theta oscillation frequency compared to ICB-S patients in the dorsal STN (depth_MER_<0.5) (Figure 1b, Supplementary Table 2). This suggests that the differences observed between ICB-R and ICB-S patients in the STN are primarily due to the dorsal region of this nucleus.

### 3.3 The Discrepancy Between ICB-R and ICB-S Groups in Neural Activity Peaks Above the Stimulation Target Region

We further investigated the localization of the differences between ICB-R and ICB-S patients. To achieve this, we divided the normalized MER depths into 5 regions (Figure 1c). We then repeated the same statistical tests described above between the two groups separately for the neurons in each region. The firing regularity metric stayed comparable across all STN regions between the two groups apart from the second region (R2) which is just above the stimulation target region (R3) (*p*<0.025) (Figure 1d, Table 3). The proportion of tonic ICB-R neurons (41.3%) was significantly higher than that of ICB-S (16.0%) (*p*=0.036, Fisher exact test) in the R2. Inversely, the ratio of bursting neurons was significantly higher in ICB-S (40.0%) compared to ICB-R (15.2%) patients in the same region. Coherently, firing regularity and coefficient of variation (cv) features were significantly different between the two groups (Table 3). A significantly heightened amplitude in the beta band was present for ICB-S compared to ICB-R groups in the R2 and R3 (Figure 1e, Table 3). Additionally, ICB-R patients exhibited longer burst durations (*p*=0.007) (Table 3) in the R3.

**Table 3.** Spatial Characteristic of Subthalamic Single-Unit Activity Between ICB-R vs ICB-S Patients. The subthalamic nucleus was divided into five regions based on normalized depths of MER recordings. The stimulation target region in both groups corresponds to the 0.4<depth_MER_<0.6 region. In each region, a statistical comparison is carried out with either the Mann-Whitney U test with the Holm-Bonferroni correction or the Fisher Exact test between ICB-R and ICB-S patients.

|  | region 1<br>(N=71) | region 2<br>(N=71) | region 3<br>(N=59) | region 4<br>(N=66) | region 5<br>(N=45) |
| --- | --- | --- | --- | --- | --- |
| **Mann-Whitney U Test** |  |  |  |  |  |
| Coefficient of Variation | 0.777 | **0.041** | 0.297 | 0.867 | 0.810 |
| Firing Regularity | 0.865 | **0.025** | 0.447 | 0.240 | 0.486 |
| Firing Rate (spikes/sec) | 0.477 | 0.119 | 0.772 | 0.708 | 0.576 |
| Theta Oscillation Frequency (Hz) | 0.088 | 0.078 | 0.621 | 0.675 | 0.484 |
| Theta Oscillation Amplitude | 0.757 | **0.046** | 0.070 | 0.426 | 0.935 |
| Beta Oscillation Frequency (Hz) | 0.206 | 0.894 | 0.375 | 0.861 | 0.855 |
| Beta Oscillation Amplitude | 0.391 | **0.001** | **0.041** | 0.784 | 0.739 |
| Burst Frequency (Hz) | 0.270 | 0.463 | 0.196 | 0.301 | 0.110 |
| Burst Duration (sec) | 0.457 | 0.159 | **0.007** | 0.708 | 1.000 |
| Inter-Burst Interval (sec) | 0.193 | 0.744 | 0.956 | 0.306 | 0.918 |
| **Fisher Exact Test** |  |  |  |  |  |
| Tonic Neurons (%) | 0.756 | **0.036** | 0.506 | 0.015 | 0.770 |
| Irregular Neurons (%) | 1.000 | 1.000 | 1.000 | 0.453 | 0.755 |
| Bursting Neurons (%) | 0.778 | **0.039** | 0.421 | 0.422 | 0.473 |
| Theta Band Oscillatory (%) | 0.582 | 0.159 | 0.754 | 0.572 | 0.745 |
| Beta Band Oscillatory (%) | 0.193 | 0.744 | 0.783 | 0.051 | 1.000 |

Based on these findings, the R2 region (0.2<depth_MER_<0.4) of STN might be the optimal region of interest (ROI) to estimate the postoperative impulsivity improvement due to notable differences observed in firing regularity, neural busts, and the amplitude of neural oscillations between two groups.

### 3.4 Neural Decoding

Given the differences described above, we investigated whether it was possible to discriminate single neurons belonging to patients in the ICB-R and ICB-S groups based on their activity (see Methods). Our focus was primarily on R2 and R3 and the corresponding relevant features in these regions (Table 3) to train and test SVM models. The classification of subthalamic neurons in R3 resulted in a balanced accuracy score of 0.639±0.159 (mean±std) and a weighted AUC score of 0.697±0.142 across five-fold in cross-validation. Higher performance was achieved when comparing the neurons of ICB patients within R2, with balanced accuracy and weighted AUC scores of 0.699±0.124 and 0.754±0.139, respectively (Figure 1f). In both cases, the trained models significantly outperformed the upper boundary of 95% percentile of random chance performances.

### 3.5 Prediction of Impulsivity Improvement

We finally attempted to define a set of neural biomarkers predicting the improvement of ICB symptoms following STN-DBS surgery based on the recordings acquired during the surgery. The cv, firing regularity, oscillation amplitudes in beta and theta bands, and the proportion of tonic and bursting neurons were selected to build multiple LDAs to discriminate between ICB-S and ICB-R patients, given the observed significant effects at the ROI R2 (Table 3).

We discovered that the combination of weak oscillations observed in both theta and beta bands at the R2 region was a strong indication for ICB recovery following STN-DBS (Figure 1g). By combining these two features, we achieved an accuracy of 9/10 under the following conditions for recovery:

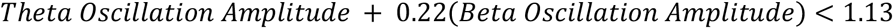

The success of the remaining two linear discriminants was limited, and a considerable number of patients fell within the margins of hyperplanes around decision boundaries, indicating a weaker degree of separation between the two groups.

### 3.6 Effect of Stimulation Targeting and Parameters on Impulsivity Improvement

Several studies reported a significant association between the STN stimulation contact side and the postoperative conditions of impulsive behaviors.^13,16^ In particular, an increasing number of publications also highlighted the role of ventral STN on impulsivity.^19,31^ To investigate these spatial aspects, we checked the normalized target positions^19^ within the STN. The median values in the dorsoventral axis were z=0.44 for both the ICB-S and ICB-R groups and the statistical comparison did not yield a significant result (*p*=1.0, Mann-Whitney U Test).

Regarding DBS stimulation parameters, no differences were observed between the ICB-R and ICB-S groups during the first-year evaluation. Stimulation frequency (130Hz) and pulse width (60 µsec) remained consistent for all patients. Despite a higher median stimulation amplitude for the ICB-S group compared to ICB-R patients in both hemispheres (right STN: 3.3V for ICB-S and 2.9V for ICB-R, left STN: 3.4V for ICB-S and 2.8V for ICB-R), the disparity was not statistically significant (*p*=0.74, Mann-Whitney U Test).

### 3.7 Correlation of Clinical and Neural Features Predicting Postoperative Recovery

Lastly, we aimed to explore any potential relationship between neural features exhibiting significant differences between ICB-R and ICB-S groups at ROI and the preoperative impulsive characteristics across all ICB patients, involving both groups.

Interestingly we noticed a significant association between the amplitude of beta oscillations at ROI and the first and second-order factors of the Barrat Impulsiveness Scale (BISFOF and BISSOF). Increased preoperative BISFOF and BISSOF motor scores indicated significantly heightened beta oscillation amplitude at ROI for all ICB patients (r^*2*^=0.739 and r^*2*^=0.759, *p*=0.015 and 0.011, Wald Test with t-distribution). Additionally, spiking regularity metrics at ROI tended to show significant correlations with preoperative motor symptoms for the ICB patients. (Supplementary Table 3). We did not observe any significant association between neural activity in this region and preoperative DRT.

## 4. Discussion

We showed that impulsivity improvement following STN-DBS surgery in PD patients is associated with specific features of neural activity in the dorsal part of the STN. In dorsal STN, just above the stimulation target region (R3), increased spiking regularity, decreased neural bursts, and weaker beta and theta oscillations serve as strong indicators for impulsivity improvement.

The relationship between the activity of single neurons in the STN has been investigated so far in a very limited set of studies. First, the processing of potential rewarding and punitive behavioral tasks at the level of single neurons in the STN was described by observing their fluctuations in the firing rate.^32^ The authors observed that PD patients with ICB showed a significantly higher rate of subthalamic neurons increasing and decreasing their firing rate under prospective reward and loss conditions, respectively, compared to non-ICB patients. Interestingly, we found that the ICB-R patients showed significantly higher firing rates in the resting state compared to non-ICB patients in the entire STN, whereas ICB-S and non-ICB PD patients remained comparable (Table 2). Later, our group compared single neuron activity recorded from explorative MER in ICB and non-ICB patients.^19^ Spiking regularity was found to be significantly higher in ventral STN for the ICB group.

However, the link between STN electrophysiology and the course of impulsive behavior following subthalamic stimulation for PD patients was never investigated before this work. Previous research indicates a strong connection between the amplitude and duration of beta bursts and the severity of motor symptoms in PD.^22,33^ We observed that the strength of beta oscillations, particularly in the dorsal STN close to the stimulation target region, is also informative for a behavioral phenomenon: the evolution of impulsive behaviors in PD. This emphasizes the significance of this hallmark subthalamic phenomenon not only in the motor domain but also in the broader context involving the behavioral aspects of PD pathology. Additionally, we demonstrated that increased spiking regularity and decreased burstiness in different segments of STN can indicate either the presence of ICB^19^ or improvement from ICB in PD patients following STN-DBS.

We observed that the degree of preoperative impulsive behaviors is strongly correlated with the strength of beta oscillations at ROI, but they did not discriminate between ICB-R and ICB-S groups. The impact of preoperative motor conditions on the post-DBS ICB outcome has already been investigated but with contradictory results. Two recent studies reported no substantial impact^8,18^, while another study reported that patients with less severe motor symptoms preoperatively are more likely to experience improvement in impulsive behaviors following the surgical intervention.^10^ The long-term use of dopaminergic medications can induce impulsive behavior and DBS-induced impulsivity recovery might be induced by the ensuing decrease in medication intake.^4,9–12^ In the scope of our study, we compared ICB-R and ICB-S groups showing comparable preoperative and postoperative motor symptomology and pharmacological treatment to not introduce confounding factors to neural analyses (Table 1). Hence, we did not observe the aforementioned differences in our cohort groups due to our experimental design. In contrast to previous studies^18^, our data did not demonstrate a significant association between preoperative impulsivity profiles and postoperative ICB recovery. The limited size of our dataset may have contributed to the lack of significant findings regarding this aspect.

While the primary limitation of our study was indeed the relatively small number of patients included with only 10 patients with ICBs, we implemented multiple robustness analyses, as described in the methods section, to mitigate the impact of this bias. Expanding the cohort size within both ICB groups would undoubtedly enhance the generalizability of our findings across the PD population exhibiting impulsive behaviors. Nevertheless, we believe that our systematic approach has provided compelling evidence supporting the involvement of subthalamic beta oscillations not only in the motor but also in the behavioral domain of PD pathology.

## 5. Conclusion

We have shown that low-frequency oscillations and spiking regularity of STN neurons extracted from standard explorative MER, particularly the region above the stimulation target in dorsal STN, could support an accurate prediction of postoperative impulsivity improvement in PD patients following STN-DBS surgery.

## 6. Acknowledgements

We express our sincere gratitude to Mr. Angel H. Cedric for his wisdom throughout this research.

## 9. Data Availability Statement

The data supporting the findings of this study are available on request from the corresponding author. The data is not publicly available due to privacy or ethical restrictions.

## 10. Author Roles

AK, LC, MV, FM, FT, and AM contributed to the conception and design of the study, data analysis, interpretation, drafting, and revising of the article. SR, LC, AG, SV, FP, DG, SM, and GG contributed to the conception and design of the study, data collection, interpretation, and revising of the article.

