## Supplementary Material for "Low-Frequency Activity in Dorsal Subthalamic Nucleus Predicts Impulsivity Improvement Following Deep Brain Stimulation in Parkinsonian Patients"

**Supplementary Table 1. Postoperative Clinical Characteristic, and Dopaminergic Replacement Therapy Comparison Between ICB-R, and ICB-S Patients.** The values presented in the table indicate the median values. The statistical comparison is made with the Mann-Whitney U test with the Holm-Bonferroni correction.

|  | Evaluation_postoperative_ | | | Evaluation_postoperative_ - Evaluation_preoperative_ | | |
| --- | --- | --- | --- | --- | --- | --- |
|  | **ICB-R** | **ICB-S** | **p-value** | **ICB-R** | **ICB-S** | **p-value** |
| Hoehn-Yahr Drug Off | 2.00 | 2.00 | 0.831 | -0.5 | -0.5 | 1.000 |
| Hoehn-Yahr Drug On | 1.00 | 2.00 | 0.734 | 0.0 | 0.0 | 1.000 |
| Hoehn-Yahr On-Off Difference | -0.50 | -0.50 | 0.664 | 0.5 | 1.0 | 1.000 |
| UPDRS III Drug Off | 8.00 | 15.00 | 0.091 | -10.0 | -11.0 | 0.833 |
| UPDRS III Drug On | 5.00 | 7.00 | 0.237 | 6.0 | -1.0 | 0.600 |
| UPDRS III On-Off Difference | -3.00 | -10.00 | 0.203 | 11.0 | 13.0 | 0.917 |
| Levodopa Dose | 705.00 | 965.00 | 0.463 | -395.0 | -58.0 | 0.421 |
| LEDD | 105.00 | 160.00 | 0.421 | -495.0 | -310.0 | 0.421 |
| DA LEDD | 600.00 | 650.00 | 1.000 | -100.0 | -240.0 | 0.600 |

**Supplementary Table 2. Statistical Comparison between ICB-R vs ICB-S Patients in Ventral and Dorsal STN.** The subthalamic nucleus was divided into dorsal (normalized depth<0.5) and ventral (normalized depth≥0.5) regions. The values presented in the table indicate the median values. A statistical comparison is carried out with either the Mann-Whitney U test with the Holm-Bonferroni correction or the Fisher Exact test between ICB-R and ICB-S patients in both regions. ******* *P*<0.001

|  | Dorsal STN | | | | Ventral STN | | |
| --- | --- | --- | --- | --- | --- | --- | --- |
|  | **ICB-R**  **(N=84)** | **ICB-S**  **(N=59)** | | **p-value** | **ICB-R**  **(N=111)** | **ICB-S**  **(N=58)** | **p-value** |
| Mann-Whitney U Test | | | | | | | |
| Coefficient of Variation | 1.160 | | 1.245 | 0.055 | 1.172 | 1.146 | 0.593 |
| Firing Regularity | 0.091 | | -0.019 | 0.059 | 0.141 | 0.214 | 0.185 |
| Firing Rate (spikes/sec) | 14.377 | | 12.404 | 0.422 | 14.164 | 11.897 | 0.782 |
| Theta Oscillation Frequency (Hz) | 6.000 | | 5.000 | **0.014** | 6.000 | 5.000 | 0.440 |
| Theta Oscillation Amplitude | 0.319 | | 0.444 | 0.089 | 0.255 | 0.348 | 0.869 |
| Beta Oscillation Frequency (Hz) | 20.000 | | 19.000 | 0.269 | 23.000 | 24.000 | 0.285 |
| Beta Oscillation Amplitude | 0.423 | | 0.619 | ******* | 0.324 | 0.335 | 0.586 |
| Burst Frequency (Hz) | 148.936 | | 154.544 | 0.488 | 142.894 | 118.168 | 0.080 |
| Burst Duration (sec) | 0.065 | | 0.060 | 0.365 | 0.101 | 0.069 | 0.110 |
| Inter-Burst Interval (sec) | 1.207 | | 0.763 | 0.205 | 1.214 | 0.830 | 0.522 |
| Fisher Exact Test | | | | | | | |
| Tonic Neurons (%) | 29.730 | | 15.517 | 0.060 | 28.571 | 42.373 | 0.108 |
| Irregular Neurons (%) | 45.946 | | 46.552 | 1.000 | 38.095 | 33.898 | 0.724 |
| Bursting Neurons (%) | 24.324 | | 37.931 | 0.075 | 33.333 | 23.729 | 0.264 |
| Theta Band Oscillatory (%) | 20.721 | | 29.310 | 0.254 | 30.952 | 22.034 | 0.259 |
| Beta Band Oscillatory (%) | 33.333 | | 44.828 | 0.180 | 30.952 | 18.644 | 0.122 |

**Supplementary Table 3. The correlation analysis between neural features with preoperative clinical features for ICB patients.** Only neurons from the ROI were included in the analysis. Median values of neural features exhibiting statistically significant differences between ICB-R and ICB-S groups were measured for each patient. Spearman’s correlation between these neural features with preoperative clinical features was assessed. The significance of the correlation coefficient was determined using the Wald Test with t-distribution. Only significant results were given.

| clinical feature | neural feature | correlation | p-value |
| --- | --- | --- | --- |
| UPDRS III Drug Off | Coefficient of Variation | 0.659 | 0.038 |
| Hoehn-Yahr Drug Off | Firing Regularity | -0.806 | 0.005 |
| UPDRS III Drug Off | Firing Regularity | -0.744 | 0.014 |
| UPDRS III Drug Off | Tonic Neurons (%) | -0.802 | 0.005 |
| BISFOF Motor | Beta Oscillation Amplitude | 0.739 | 0.015 |
| BISFOF Cognitive Instability | Beta Oscillation Amplitude | 0.871 | 0.001 |
| BISSOF Attention | Beta Oscillation Amplitude | 0.798 | 0.006 |
| BISFOF Motor | Beta Oscillation Amplitude | 0.759 | 0.011 |
| BISSOF Total | Beta Oscillation Amplitude | 0.772 | 0.008 |
| BISSOF Total | Theta Oscillation Amplitude | 0.662 | 0.037 |
